# Left Bundle Branch Pacing Facilitated by Novel Surface Electrocardiography in Comparison with Electrophysiology Recording System

**DOI:** 10.1101/2023.05.22.23290368

**Authors:** Lan Su, Ling Zhu, Songjie Wang, Shengjie Wu, Xiao Chen, Zhouqing Huang, Liangping Wang, Lei Xu, Xiaohong Zhou, Weijian Huang

## Abstract

**Background:** Left bundle branch pacing (LBBP) had been proven to be feasible and safe in patients with a pacemaker indication. In this study, we assessed the feasibility and safety of LBBP procedure using simplified ECG monitoring and LBBP criteria in comparison with that by using the conventional EP system and currently adopted LBBP criteria.

**Methods:** The single-center study included 143 consecutive patients from March 2021 to January 2022. The operator was single-blind to the electrophysiology recording system (EP system), and only observed the electrophysiological characteristics of the four-lead ECG monitor and the pacing system analyzer (PSA) by naked eye. Other researchers kept synchronized records of the EP system, and analyzed whether the data were correct and consistent after the operation. Intraoperative data were collected and the safety of 3-month follow-up after operation were evaluated.

**Results:** Of 143 patients enrolled, 139 successfully performed LBBP, with a success rate of 97.2%, and the judgement concordance with EP system was 99.3%. The total operation time was 78.9±26.5min, the total fluoroscopy time was 9.5±6.1min, the fluoroscopy time of left bundle branch (LBB) lead deployment 3.0±2.6min, which had no significant difference with previous studies. Ventricular septal perforation occurred in 2 patients during the operation. Pacing parameters were stable and heart function improved during follow-up.

**Conclusions:** The simplified LBBP implantation method without an EP system and only relying on a simplified ECG combined with an analyzer is clinically feasible and safe and can be promoted in clinical practice.

## Introduction

Cardiac conduction system pacing activates the ventricles in a physiological sequence, including His bundle pacing (HBP) and left bundle branch pacing (LBBP)^1–3^. As HBP has limitations such as difficult operation, high and unstable pacing threshold, and low success rate^4^, Huang et al. first introduced the clinical application of LBBP in 2017, which has the advantages of low and stable pacing threshold and high success rate^5–7^. Clinical studies in recent years have verified its procedural feasibility, mid-term safety and favorable clinical benefits^8–10^. The LBBP in clinical practice has grown rapidly. Currently, most operators perform the LBBP procedure using a confiscated electrophysiology recording system (EP system) that has measuring capability of 12-lead ECG and cardiac mapping to measure changes of Stim-LVAT and observe the transformation of ECG QRS morphology and LVSP (S-LBBP) to NS-LBBP and injury of current in intracardiac electrogram (EGM) to confirm LBB capture^11–13^. However, many hospitals do not have the EP system for LBBP implantation, limiting the adoption of LBBP in routine clinical practice. Thus, the objective of the present study was to assess the feasibility and safety of LBBP procedure using simplified 4-lead body surface ECG monitoring and corresponding LBBP criteria in comparison with that by using the conventional EP system and currently adopted LBBP criteria.

## Methods

### 1. Study patients

This single-center prospective study included consecutive patients from March 2021 to January 2022, and the inclusion criteria were: (1) an indication for treatment of bradycardia or cardiac resynchronization therapy; (2) age older than 18 years; (3) signed informed consent. Exclusion criteria were: (1) ischemic cardiomyopathy; (2) nonspecific intraventricular conduction delay (IVCD); (3) ventricular septal hypertrophy (end-diastolic thickness over 15 mm); (4) pacemaker upgrade. The approval of the Ethics Committee of the first affiliated hospital of Wenzhou Medical University was obtained (KY2022-078), and informed consent was obtained from all recruited patients. The trial was conducted in accordance with the principles of the Declaration of Helsinki and was registered in ClinicalTrials.gov (NCT05553431).

### 2. Implantation process

The SelectSecure pacing lead (3830, Medtronic Inc.) and C315His delivery sheath (Medtronic Inc.) were used in all patients. The CPS Direct™ PL Peelable Outer Guide Catheter (Abbott Inc.) will be used when necessary. In order to meet the needs of operation by the simplified method, we developed a new modified ECG lead system (Figure 1) that had four-lead body surface ECG monitoring (IntelliVue MP60, Philips Inc) to simulate V5, V1, II and III respectively, the amplitude of the QRS complex was set to 1mv with a sweep speed of 50mm/s. The placement of 4-channel ECG electrodes is shown in Figure 1. Since these leads are simulated by a modified chest-limb lead method, QRS complexes are not identical to those in the standard 12-lead ECG. However, the similarity of QRS morphology between modified V1 lead and EP-system is 100%. The similarity of modified V5 lead vs. that of 12-lead ECG is mainly present in the wave direction. The pacing system analyzer (PSA, model number: 2290; Medtronic Inc.) could display and record large LBB potential and injury current through EGM and test pacing parameters. The filter default setting was 0.5-1000Hz. The Electrophysiology recording system (GE CardioLab EP Recording System 2000; GE Healthcare, Milwaukee, WI) was simultaneously used for recording 12-lead ECG and EGM, and the filter was usually set to 30-500 Hz for collecting high-frequency signals, or 0.5-500Hz when recording ventricular or LBB current of injury (COI).

**Figure 1.**
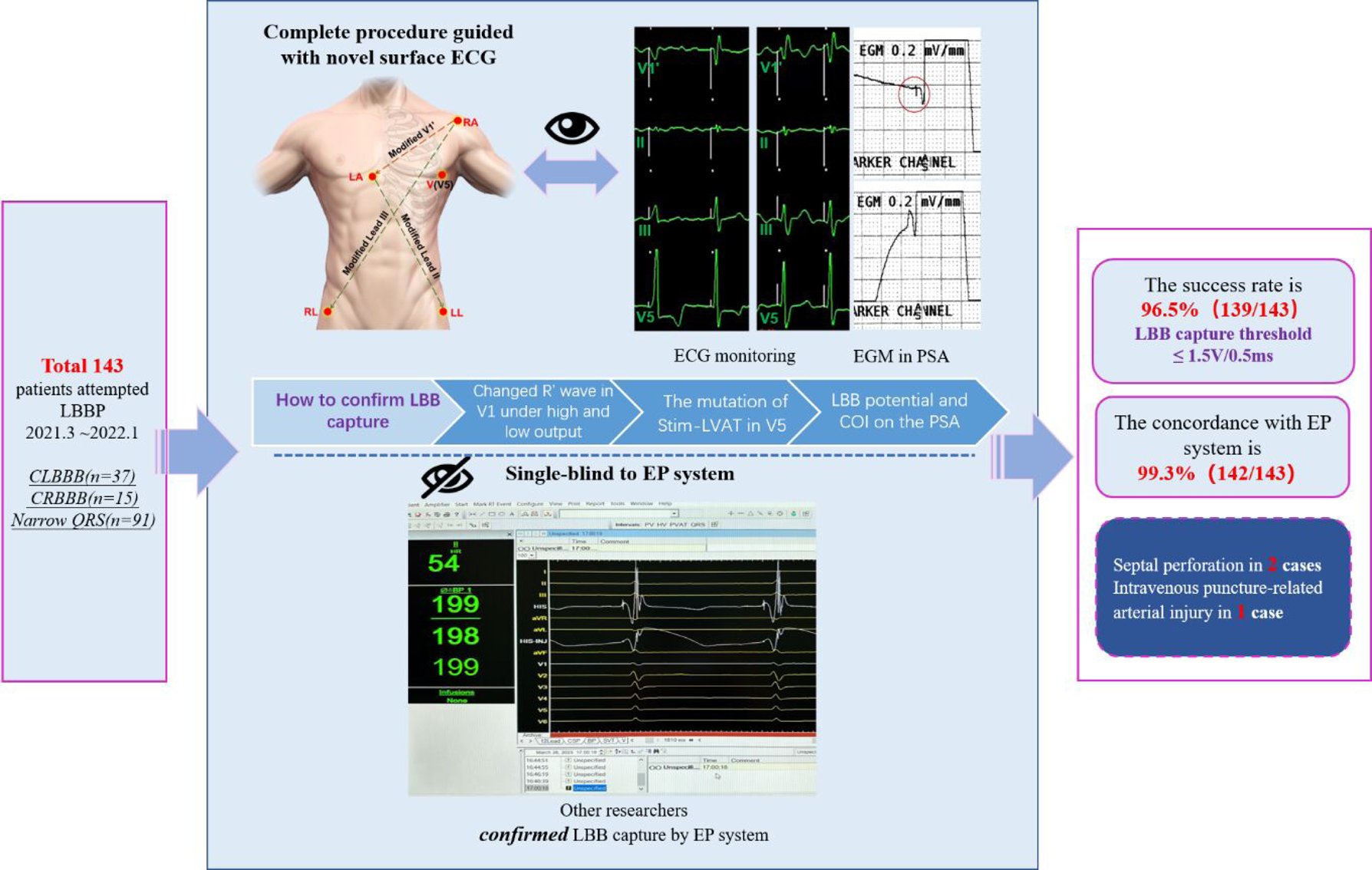
New Modified ECG Lead System. The new modified ECG lead system used a five-electrode monitoring cable with four channels. The right arm (RA) electrode was placed in left shoulder and the left arm (LA) electrode was placed in the right fourth intercostal space to obtain a modified V1’ lead configuration. The left leg (LL) electrode and the right leg (RL) electrode were placed in left and right lower limbs respectively. Modified lead II was obtained by LA and LL electrodes, while modified lead III was obtained by RA and RL electrode. The fifth(V) electrode was placed in the stand V5 lead position.

The specific method of LBBP implantation has been well described previously^11, 12^. The procedure for performing LBBP using 4-lead ECG monitor and PSA is as follows:

i. Localization of the His bundle and RV entry side for LBBP lead was conducted by a quick His mapping with fluoroscopic images and engaging the lead tip at the right ventricular septum for LBBP (Figure 2).
ii. Pacing was performed as the lead was screwed in. When PVCs with RBBB morphology or paced RBBB pattern in lead V1 occurred, it was indicated that the tip of lead was close to the LBB area. At this moment, the screwing was stopped and pacing test was conducted. Observed LBB potential or with COI on intracardiac electrocardiogram of PSA could assist determining accurate positioning of LBB.
iii. Following step ii, pacing was performed at a low output (1V/0.5ms) and then at a high output (10V/0.5ms) to observe the changes of QRS waves. Once the shape of R wave in lead V1 changed (emerging R wave, widening R’ wave and narrowed R’ wave), it was considered to capture the LBB. Gradually the pacing output was gradually reduced, starting at 1V/0.5ms each time and then 0.1V/0.5ms each time when the output was low. In the process of reducing output, the operator judged whether there was a sudden change in Stim-LVAT by the naked eye to confirm transformation of NS-LBBP, S-LBBP and LVSP in the surface ECG (Figure 3).
iv. The pacing test ended if the parameters were satisfactory; if the test parameters were unsatisfactory or LBB was not captured, the lead depth or reposition was adjusted until LBB was captured.

**Figure 2.**
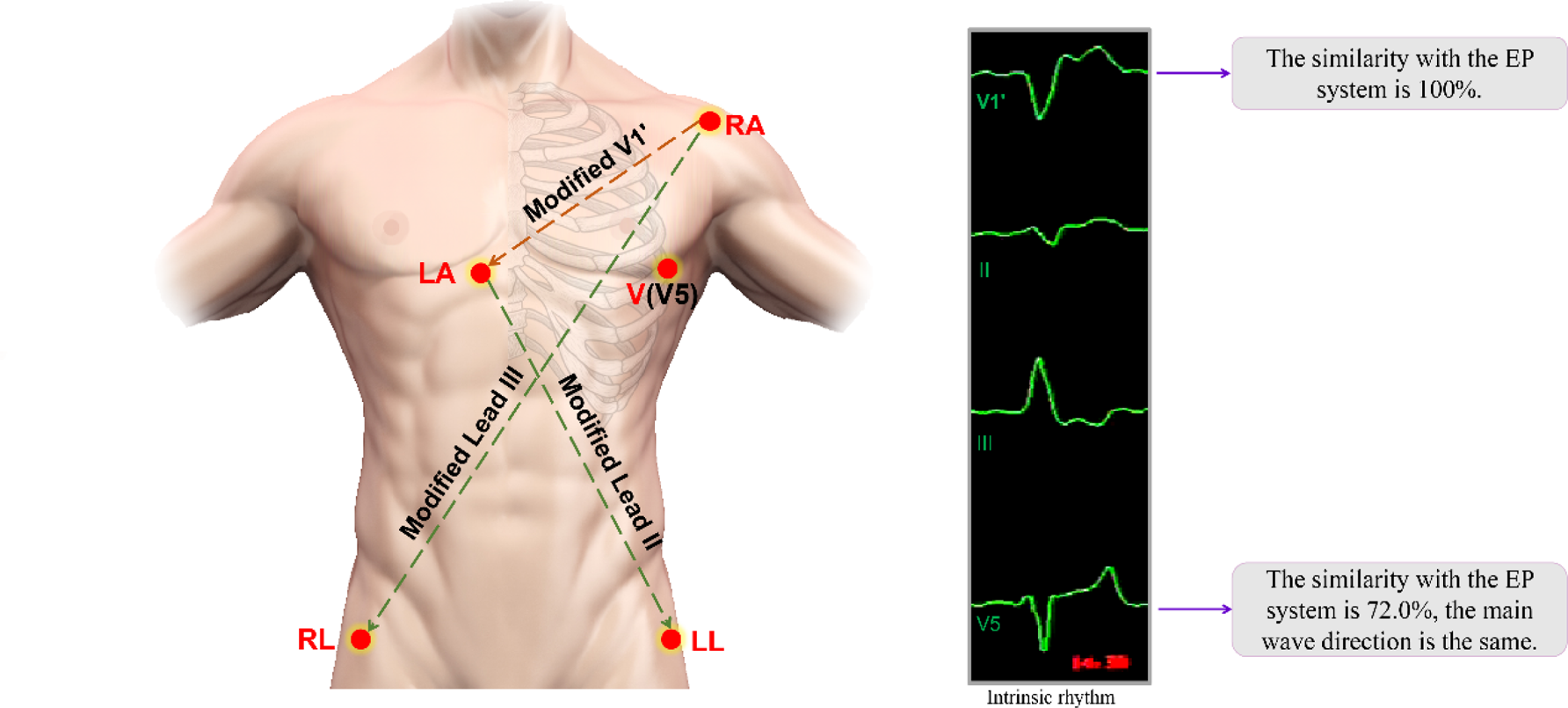
The Routine of Left Bundle Branch Pacing. The operation of LBBP had been described in detail in previous studies, and several unconventional methods of His bundle location had emerged. A simplified method using ECG monitor and PSA was proposed for pacing mapping in the LBB area. HB= his bundle; ICE= intracardiac echocardiogram; PSA= pacing system analyzer.

**Figure 3.**
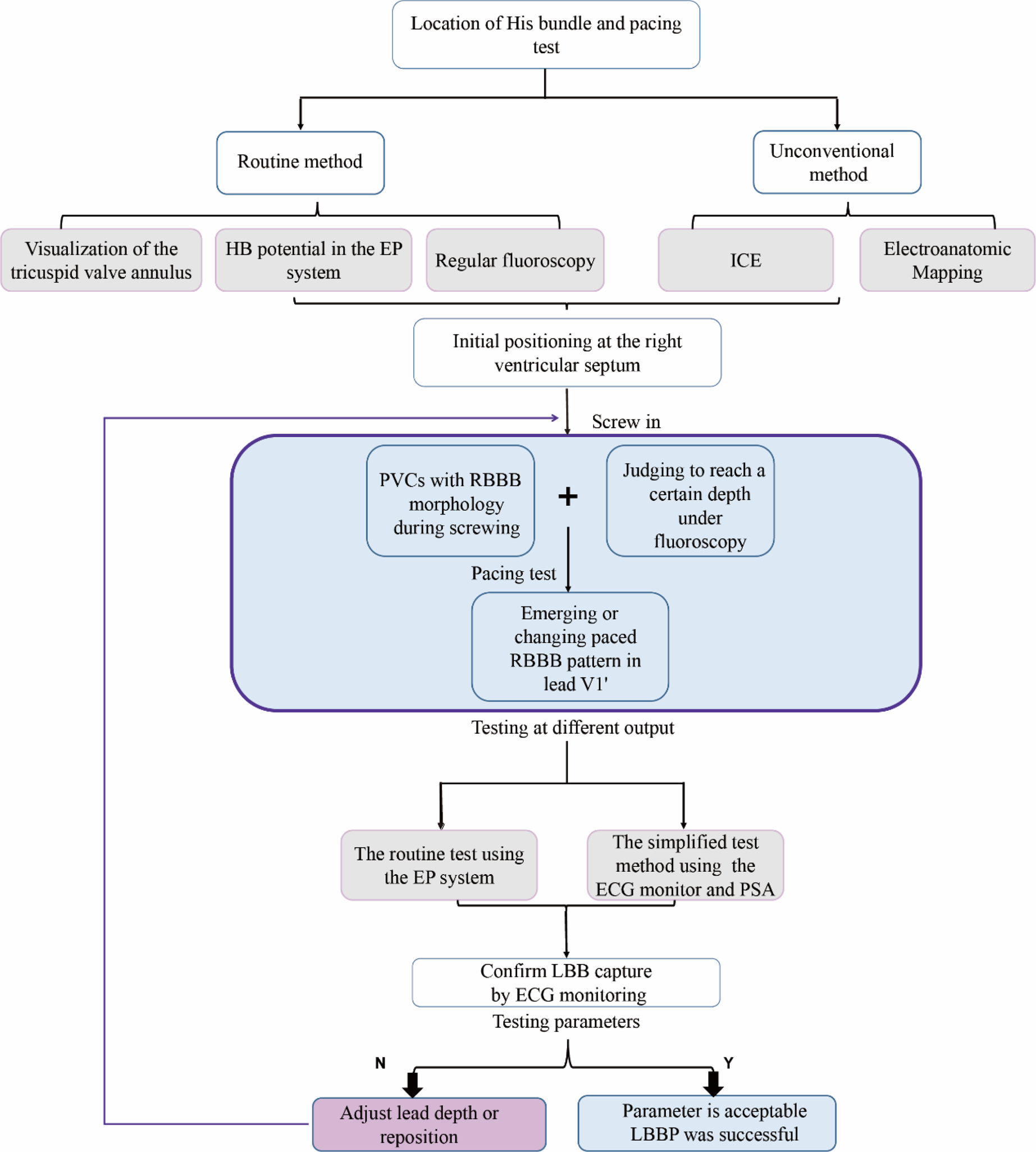
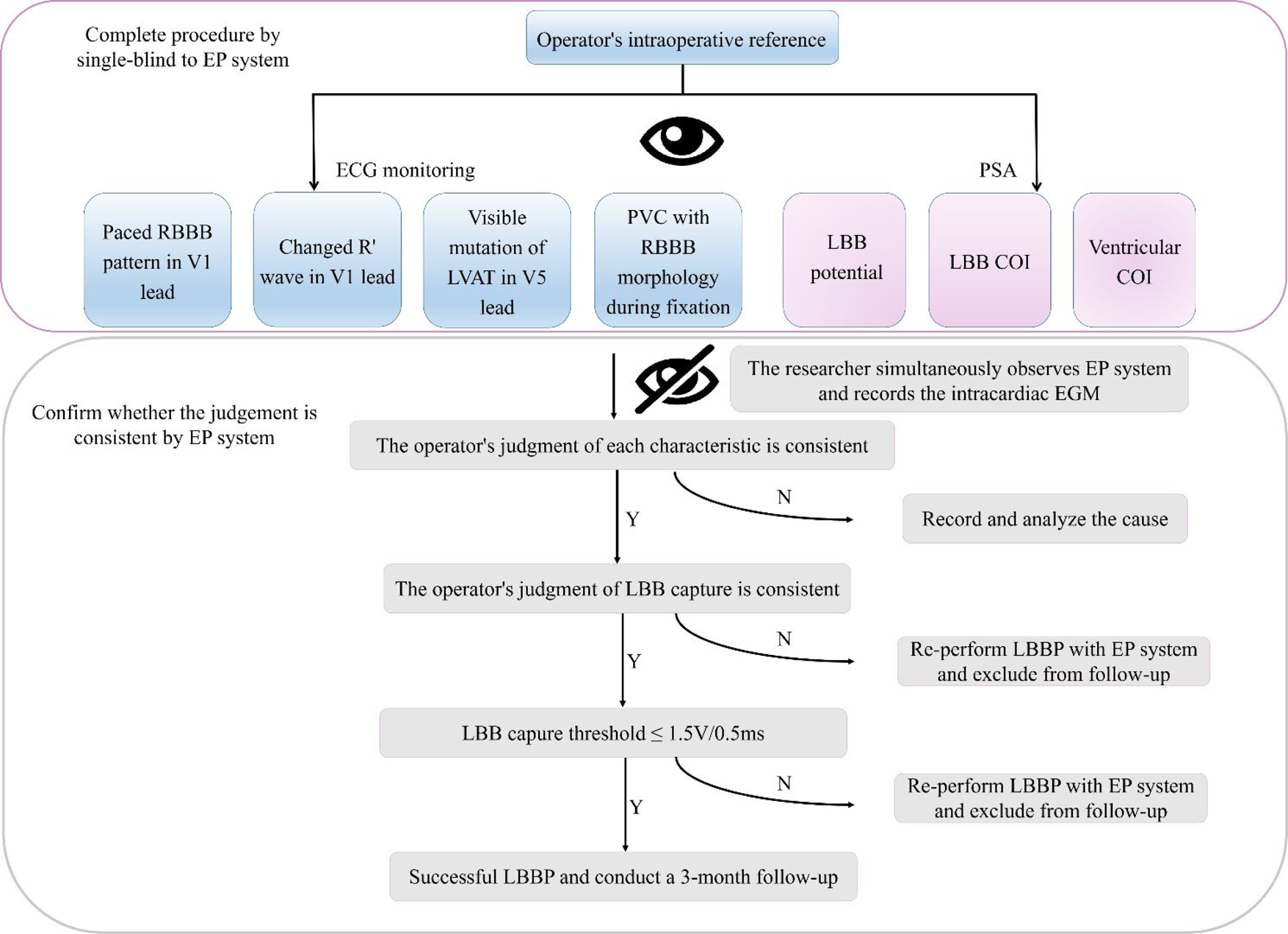
Flowchart for the Simplified Left Bundle Branch Pacing. The operator was single-blind to the EP system and determined the LBB capture by signs of ECG monitoring and PSA. Other researchers synchronously observed the EP system, evaluated the operator’s judgments, and recorded relevant information. PSA= pacing system analyzer; COI= current of injury; EGM = electrogram.

During the operation, the LBBP confirmation by the simplified 4-lead ECG and Analyzer was conducted by the operator’s visual assessment. At the same time, the operator was blinded to EP system that was conducted by an engineer with ample experience in EP measurements. The operator’s visual observation of the electrophysiological characters included the RBBB pattern of the paced QRS, the change of paced QRS wave under high and low output, the mutation of Stim-LVAT in V5 lead, PVC with RBBB morphology during fixation and LBB potential of EGM on the PSA. Simultaneously, the well-trained engineer independently recorded ECG and EGM of the EP system and measured all LBBP-related variables and confirmed LBB capture based on the current LBBP criteria^13^. At the end of the operation, the consistency between the operator’s judgement of LBBP and engineer’s LBBP confirmation was determined, and the electrophysiological characteristics of the EP system and PSA were further analyzed (Figure 3).

Intraoperative data were collected, including operation time, fluoroscopy time, fluoroscopy time of His lead placement, fluoroscopy time of LBBP lead placement, pacing parameters, intraoperative complications, and operation success rate.

### 3. Definition of successful left bundle branch pacing implantation

The operator confirmed the LBB capture with the capture threshold lower than 1.5V/0.5ms. Intra-implantation, if operator’s LBBP judgement was consistent with LBBP confirmed by the EP system, the LBBP implantation was deemed as success; if the operator’s LBBP confirmation was confirmed as non-LBBP by the EP system, the LBBP implantation was assigned as failure and the reasons of failure were truthfully documented and intraoperative information was recorded, then the operator re-performed LBBP with the assistance of the EP system (Figure 3).

### 4. Definition of selective LBBP(S-LBBP) and non-selective LBBP(NS-LBBP)

LBBP criteria have been well described in previous studies^13–15^, based on which we further clarify the LBBP criteria for the simplified ECG-guided LBBP implantation as below:

1. S-LBBP is defined as capturing only the LBB: A. When the output is reduced, the R wave in lead V1 widens while the S wave in lead V5 deepens; the paced QRS in lead V1 is M or rsR’ pattern and the QRS complex duration is wide with a notch (Figure 4A). B. There is no obvious change in Stim-LVAT judged by naked eyes. C. Discrete component is visible between the pacing stimulus and the onset of ventricular potential on the EGMs of PSA (Figure 5D).
2. NS-LBBP simultaneously captures the LBB and local myocardium during pacing, therefore, the QRS morphological changes can be observed when the output voltage is changed. A. When the output is increased, the paced QRS in lead V1 becomes QR pattern or R wave is smaller, and the S wave in V5 becomes smaller; there is no obvious change in Stim-LVAT judged by naked eyes, indicating the change from S-LBBP to NS-LBBP (Figure 4A). B. When the output is increased from a capture threshold, a new R wave or an enlarged R wave in lead V1, accompanied by a significant shortening of the Stim-LVAT visible to the naked eye, suggests a transition from LVSP to NS-LBBP. C. Simultaneous capture of the conduction bundle and local myocardium has no discrete component on the EGMs of PSA (Figure 5D).

**Figure 4.**
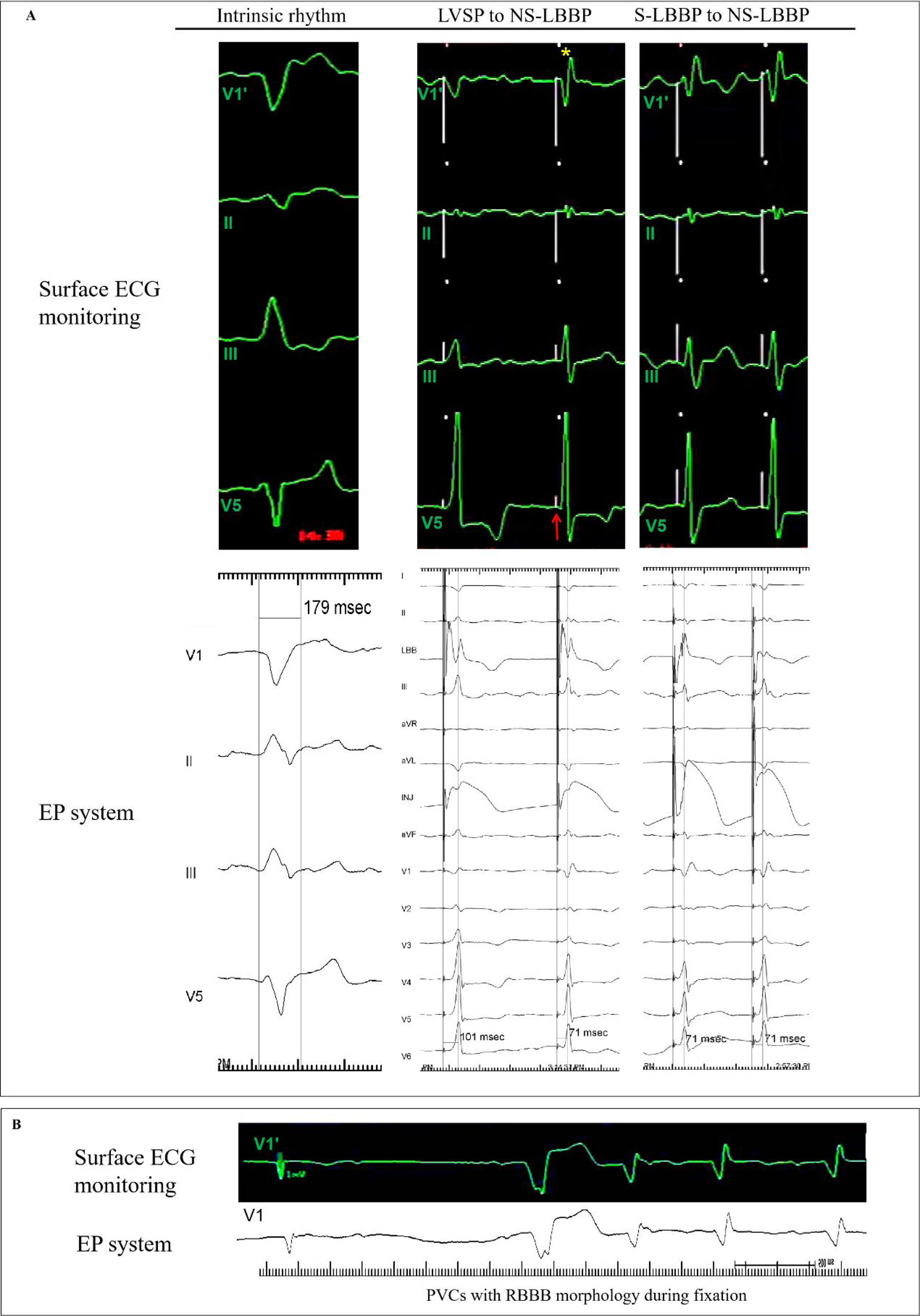
Electrophysiological Characteristics on ECG Monitoring and EP system. (A) At the initial depth, a transition from LVSP to NS-LBBP was noted as output increased, and paced RBBB pattern was observed (yellow asterisk). Stim-LVAT could be measured on the EP system and reduced from 101ms to 71ms. At the same time, it was also observed that Stim-LVAT was abruptly shortened in lead V5 on ECG monitoring (red arrow), although it was not as intuitive as EP system. At the final depth, QRS morphology shifted from S-LBBP to NS-LBBP as output increased. On ECG monitoring, it could be observed that the R wave in lead V1 of S-LBBP was wider, and the S wave in lead V5 was deeper. Stim-LVAT in lead V5 of S-LBBP and NS-LBBP were similar. (B) The PVCs with RBBB morphology were obviously different from the beat of intrinsic rhythm (the first beat), and it can be easily distinguished in ECG monitoring and EP system. LVSP= left ventricular septal pacing; NS-LBBP= nonselective left bundle branch pacing; S-LBBP= selective left bundle branch pacing; Stim-LVAT= time from stimulus to left ventricular activation; PVC= premature ventricular contraction.

**Figure 5.**
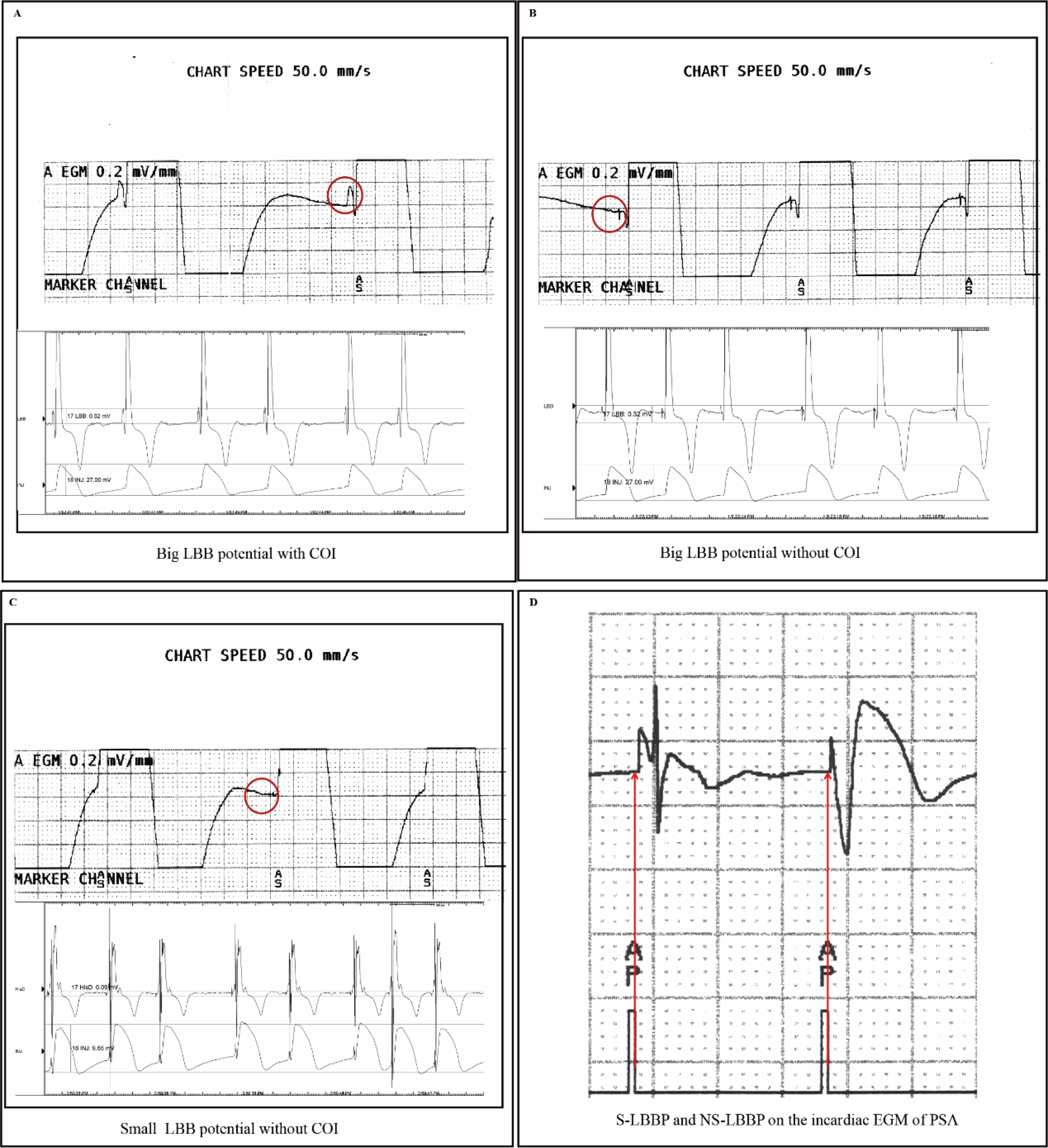
Electrophysiological Characteristics on PSA. (A, B, C) The LBB potential and COI (red circle) of three cases on PSA. One patient could be noted to have obvious LBB potential on PSA, and the amplitude of LBB potential measured on EP system was 0.2mv. And COI of ventricular and LBB potential could also be identified. (D) Obvious discrete component (red arrow) between the pacing stimulus and the onset of ventricular potential could be observed on the EGMs of PSA while S-LBBP. LBB= left bundle branch; COI= current of injury; NS-LBBP= nonselective left bundle branch pacing; S-LBBP= selective left bundle branch pacing; EGM = electrogram.

### 5. Follow-up

Patients were followed up for at least 3 months after the operation, and pacing parameters, 12-lead electrocardiogram, echocardiography, and postoperative complications were documented during follow-up.

### 6. Statistical analysis

Continuous variables are expressed as mean ± SD, enumeration data are expressed as the number of cases (%) and using χ^2^ test, measurement data using Student’s t test, paired t test to analyze the difference between baseline and follow-up, P ≤ 0.05 is considered statistically significant difference. Statistical analysis was performed using statistical software packages such as SPSS Version 23 or R Software Version 4.1.

## Results

### 1. Baseline characteristics

Patients’ baseline characteristics are summarized in Table 1. Among 143 enrolled patients, atrioventricular block was presented in 64 patients (44.8%), sick sinus syndrome was noted in 54 patients (37.8%), and 51 patients (35.7%) had atrial fibrillation. According to the QRS duration of baseline ECG, the patients were divided into CLBBB group (n=37) and non-CLBBB group (n= 106). The non-CLBBB group could be divided into CRBBB group (n= 15) and narrow QRS group (n= 91). The mean New York heart function class was 1.1 ± 1.2, and the mean New York heart function class of patients in the CLBBB subgroup was 2.3 ± 0.9, and the proportion of patients with cardiomyopathy heart failure in the CLBBB subgroup was 67.6% (25/37).

**Table 1.**
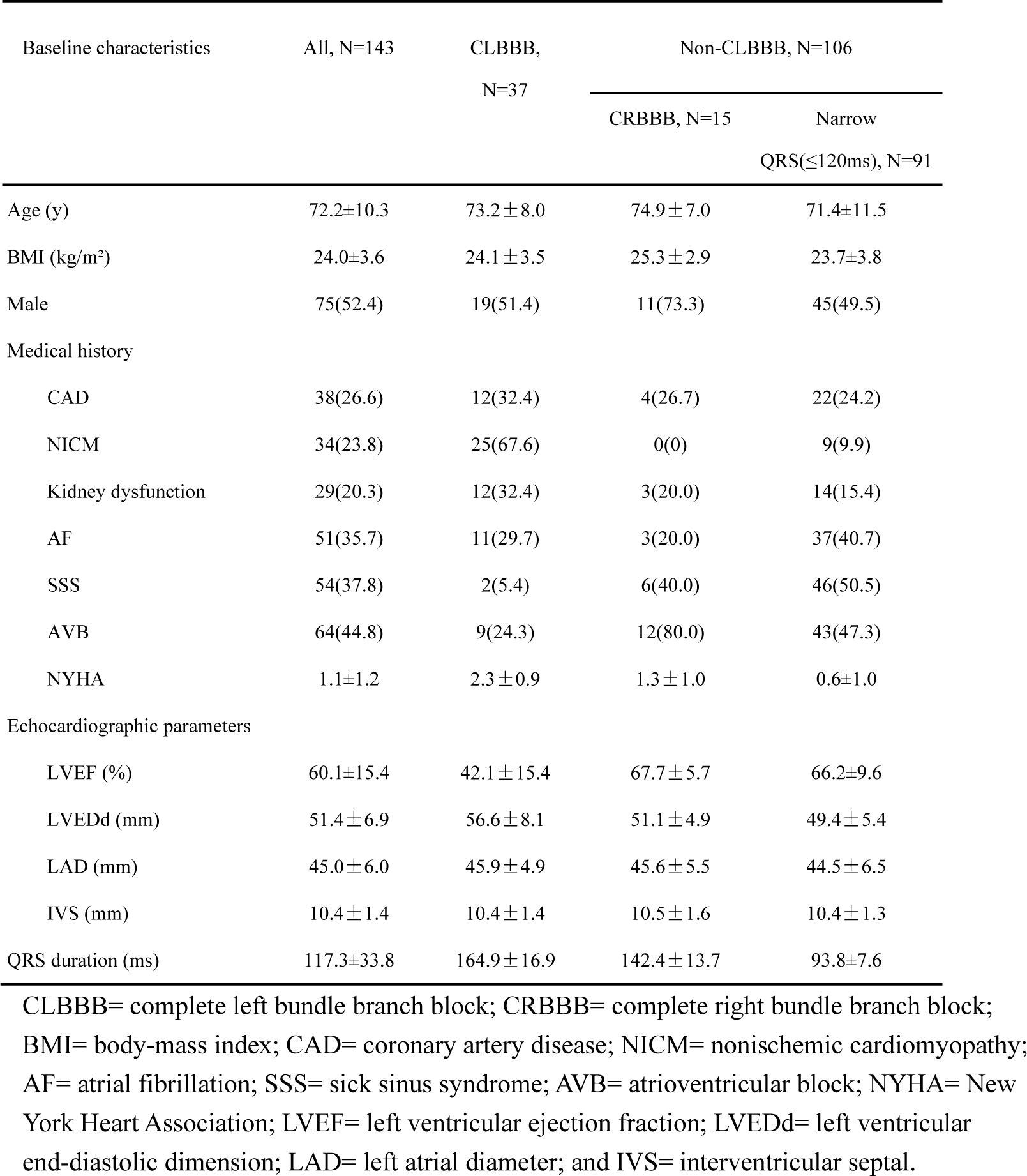
Baseline characteristics

### 2. Intraoperative characteristics

LBBP was successfully performed in 139 patients with a success rate of 97.2%, and left ventricular septal pacing was performed in the remaining 4 patients. Among the 4 patients, the pacing lead tip could not be screwed deeply because of septal fibrosis in 2 patients, high LBB capture threshold (>1.5V/0.5ms) in 2 patients. One patient was re-performed LBBP under the EP system due to inconsistent judgment, which was not included in the postoperative analysis. Total operation time (time from skin cut to suture) was 78.9±26.5min, total fluoroscopy time 9.5±6.1min and fluoroscopy time of LBB lead deployment 3.0±2.6min. (Table 2).

**Table 2.** Intraoperative Information

|  | All, N=143 | CLBBB, N=37 | Non-CLBBB, N=106 |  |
| --- | --- | --- | --- | --- |
| | | | CRBBB, N=15 | Narrow QRS($\leq 120$ms),<br>N=91 |
| Procedure duration, min | 78.9±26.5 | 91.7±22.2 | 85.9±36.3 | 72.5±24.2 |
| Total fluoroscopy time, min | 9.5±6.1 | 11.2±7.3 | 12.1±5.0 | 8.3±5.4 |
| Total fluoroscopy dose, mGy | 44.9±31.7 | 54.2±36.5 | 56.7±17.0 | 39.2±30.2 |
| Fluoroscopy time of His lead deployment, min | 0.6±0.7 | 0.7±0.7 | 0.7±0.6 | 0.6±0.7 |
| Fluoroscopy dose of His lead deployment, mGy | 3.9±3.6 | 3.7±3.4 | 3.9±2.9 | 3.9±3.8 |
| Fluoroscopy time of LBB lead deployment, min | 3.0±2.6 | 3.2±2.7 | 3.5±2.9 | 2.8±2.6 |
| Fluoroscopy dose of LBB lead deployment, mGy | 19.3±16.6 | 20.6±15.1 | 24.8±13.6 | 17.8±17.6 |
| Judgement consistent with EP system, N(%) | 142(99.3) | 37(100) | 15(15) | 90(98.9) |
| Left bundle branch pacing in final, N(%) | 139(97.2) | 37(100) | 13(86.7) | 89(97.8) |

CLBBB= complete left bundle branch block; CRBBB= complete right bundle branch block; LBB= left bundle branch; EP system= electrophysiology recording system.

### 3. Changes of surface electrocardiogram

#### (1) Paced RBBB pattern in lead V1

The operator’s ECG monitoring with simplified 4-lead ECG showed clear rSR’ or QR in RBBB pattern in lead V1 in all 144 patients, which was consistency with those observed in the EP system (Figure 4A), yielding100% accuracy. Since RBBB-shaped QRS wave can be present during left ventricular septal capture, Therefore, the paced QRS wave in RBBB pattern may not be used as an only criterion to judge the capture of the left bundle branch.

#### (2) QRS morphological changes

During the operation, unipolar pacing was performed, and the output increased from low to high. Based on ECG monitoring, the operator monitored the morphological changes of R’ wave on lead V1 of the simplified 4-lead ECG. We considered LBB capture once the change of R’ wave in lead V1 occurred during pacing output change. With the ECG monitor, the operator could observe two types of LBB capture transitions: S-LBBP to NS-LBBP or LVSP to NS-LBBP (Figure 4).

##### NS-LBBP to S-LBBP

The operator observed the changes of NS-LBBP to S-LBBP during decrement of pacing outputs in 126 patients, which was 100% consistent with observation by the EP system. Among 126 patients, 114 patients showed the widening of the R’ wave in lead V1, and all 126 patients showed the deepening of the S wave in lead V5 (Table 3).

**Table 3.**
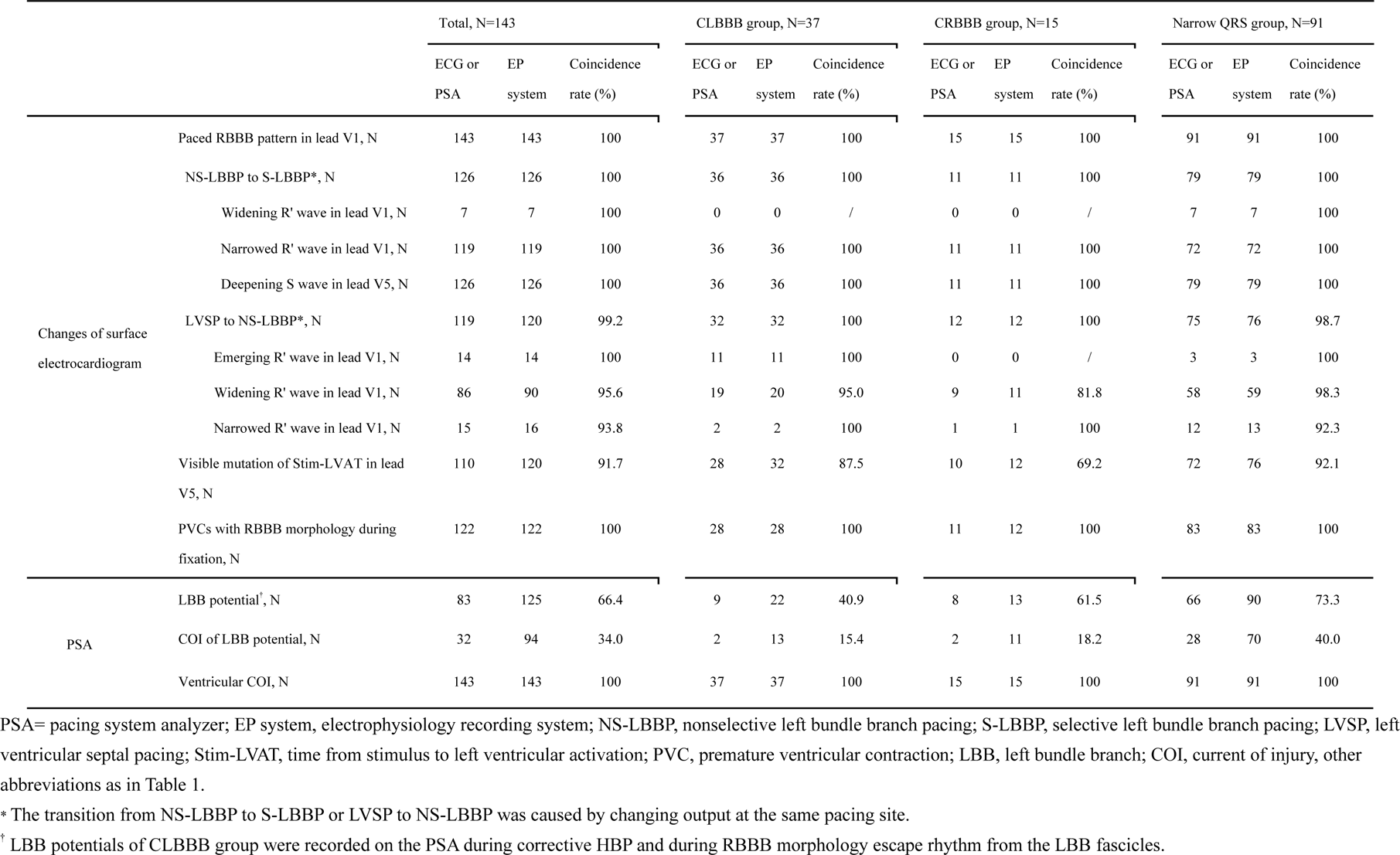
Comparison of Electrophysiological Characteristics between ECG monitoring, PSA, and EP System

##### LVSP to NS-LBBP

Changes in LVSP to NS-LBBP were observed in 121 patients on the EP system. The 121 patients showed a sudden shortening of Stim-LVAT on the V5 lead with changes of QRS wave on the V1 lead (14 cases of emerging R wave, 90 cases of widening R’ wave and 17 cases of narrowed R’ wave). The emerging R wave meant that the QRS morphology changed from QS type to QR/rSR’ type. The operator made a successful judgment of 120 patients through the four-lead ECG monitoring, and only 1 patient was wrongly judged during the operation, which was inconsistent with the record of the EP system. This case had second-degree type II atrioventricular block with a QRS duration of 100ms. When the output was increased, the QRS pattern changed from LVSP (Stim-LVAT=78ms) to NS-LBBP (67ms), and LVSP was misjudged as S-LBBP.

Notably, the phenomenon of the emerging R wave mainly occurred in the CLBBB group. And there were 17 patients with reduced R waves when LVSP to NS-LBBP. We considered it to be caused by retrograde activation of the right bundle branch during non-selective capture of LBB^16^.

#### (3) Visible mutation of Stim-LVAT in V5 lead

The operator could clearly observe the visual changes of Stim-LVAT(19.4±8.4ms) in 110 patients on ECG monitoring while the EP system detected changes of Stim-LVAT in 120 patients (18.7±8.4ms), yielding a 91.7% concordance. In patients whose Stim-LVAT was not clearly judged to be shortened, the operator judged the LBB capture by the change in the QRS morphology. There was no statistical difference between CLBBB group (87.5%, N=28/32), CRBBB group (69.2%, N=10/12) and narrow QRS group (92.1%, N=72/76). Notably, in patients (93/120) with a Δ Stim-LVAT ≥ 12ms, the change could be recognized by the naked eye with a specificity of 100%.

#### (4) PVC with RBBB morphology during fixation

When the tip of the lead is close to the LBB area, mechanical stimulation of the LBB or the adjacent septal myocardium during screwing-in can lead to PVC with RBBB pattern in ECG lead V1 (Figure 4B). PVC with RBBB pattern during lead screw-in attempt was observed in 122 of 144 patients (84.7%) with 100% consistency between the operator’s observation and the EP system. (Table 3).

### 4. Phenomenon of intracardiac electrocardiogram

LBB potentials were recorded on EP system in 86.8% patients (125/144). In the narrow QRS group, the operator only observed the LBB potential in the EGM of the PSA in 73.3% of patients (66/90), which was much lower than the potential recording rate by the EP system. The reason might be that PSA could not adjust the filtering of the recording, so some P potentials with smaller amplitudes were ignored (Figure 5). When the amplitude of LBB potential of non-CLBBB group on the intracardiac EGM of the EP system was greater than or equal to 0.16mv (N=41), the sensitivity of PSA to detect LBB potential was 100%. The amplitude of the LBB potential that could be recorded by PSA was 0.28±0.26mV, while the amplitude of the LBB potential that could be recorded on the EP system but not on PSA was 0.08±0.04mV. Potential-related COI in the PSA EGM was also observed in 38.8% (56/144, or 67.5% of 83 patients with LBB potential) patients. The EGM ventricular COI recording rate in the PSA reached 100% (144/144), which was consistent with EP system (Table 3).

### 5. Pacing parameters

Total 139 patients were judged to successfully receive LBBP. One patient did not complete the follow-up, the follow-up completion rate was 99.3%. One patient was re-performed LBBP using EP system and 2 patients had the pacing capture threshold of the LBB increased to more than 3V/0.5ms during follow-up, were not included in the statistical analysis. Finally, the pacing parameters of 135 patients were finally included in the statistical analysis. The LBB capture threshold was slightly increased at 3-month follow-up (0.60±0.25V/0.5ms) compared with the immediately postoperative period (0.53±0.23V/0.5ms, P=0.008). There was no statistically significant change in LV septal myocardium capture threshold between 3-month follow-up (0.63±0.25V/0.5ms) and the immediately postoperative period (0.60±0.26V/0.5ms, P=0.170), the unipolar impedance was significantly lower at 3-month follow-up (498.9±108.5 Ω) than the immediate postoperative period (377.2±47.4Ω, P<0.001).

In this study, it was found that patients with LBB potential accompanied by potential COI during implantation had a lower intraoperative capture threshold than patients without LBB potential COI (0.74 ± 0.32 versus 0.81 ± 0.40V/0.5ms, P=0.463), and further decreased on the first day after operation (0.43±0.18 versus 0.50±0.30V/0.5ms, P=0.157), and remained low during the 3-month follow-up (0.60 ± 0.24 versus 0.63± 0.22V/0.5ms, P=0.603), but there was no statistical difference.

### 6. Intraoperative and postoperative complications

During the operation, 2 patients had ventricular septal perforation, which showed that the EGM amplitude on the PSA became smaller, the impedance decreased below 450Ω from 600Ω, and the capture threshold increased from less than 1.5V/0.5ms to more than 5V/0.5ms. The pacing lead were repositioned. In 2 patients, the LBB capture threshold increased due to the sheath was withdrawn due to the micro-displacement of the lead tip, and were resolved after changing the position and re-fixation. One patient experienced an injured small branch of thoracoacromial artery during left vein path puncture, which was recovered without intervention. There were no serious complications occurred during follow-up.

### 7. Cardiac function and LVEF improvement

Three groups of NYHA functional class improved compared with the baseline, and the CLBBB group had the most significant improvement (1.6±0.7 from the baseline 2.3±0.9, P<0.001). Three-month follow-up in the CLBBB group showed a significant improvement in LVEF compared with baseline (53.7±11.9% versus baseline 41.3±14.7%, P<0.001), and there was no statistical difference between the CRBBB group (66.7±5.1% versus 68.0±6.3%, P=0.548) and the narrow QRS group (66.7±9.1% versus 65.9±8.6%, P=0.396).

## Discussion

### 1. Feasibility and safety

In this study, without the guidance of an EP system, the operator could rely on the simplified 4-lead body surface ECG and the PSA to accurately determine whether LBB was captured during LBBP operation with a 99.3% concordance between the operator’s visual judgement of LBB capture versus the confirmation by the EP system. Only one patient with narrow QRS failed to be correctly confirmed by both the operator and the EP system. Compared with our previous studies, the intraoperative operation time was similar (78.8±26.4 versus 86.4±43.5min in the previous report^9^), and the fluoroscopy time did not increase compared with previous large-scale studies (9.5±6.1min versus 9.7±7.3min reported in the previous studies^17^). Our findings confirm the feasibility of LBBP operation without a need of an EP system. During follow-up, 2 patients with elevated capture thresholds still maintained low LV septal capture thresholds (0.5V/0.5ms and 0.75V/0.5ms), which ensured the safety of pacing. There were no serious complications during the operation, and the LBB capture threshold remained low and stabilized at the 3-month follow-up, showing a good safety consistent with previous studies of LBBP under standard practice.

### 2. The main point of the simplified method

The main work of LBBP confirmation utilizing the simplified method under a 4-lead ECG monitoring alone is to observe the changes of the 4-lead ECG on the body surface, including: ① judge the QRS morphological changes of LVSP, NS-LBBP and S-LBBP should be judged under high and low voltage output; ② confirm LBB capture with the shortening of Stim-LVAT in V5 lead that can be discerned by the naked eye; ③ observe LBB potential or COI in the intracardiac electrocardiogram of PSA to confirm LBB location. If only relying on one of the above indicators, it is difficult for operator to accurately determine the capture of LBB, however, with the above two indicators the accuracy of the diagnosis of LBB capture can be significantly improved. All patients in the present study had pacing patterns in the form of RBBB in ECG. And the patient’s ordinary ECG showed the change of the QRS shape, with the visible shortening of Stim-LVAT, the combination of the two can complete the accurate judgment of high concordance with to the LBBP confirmation by the EP system. With the recording of LBB potential and COI in the intracardiac electrocardiogram of PSA, the capture of the LBB can be completed in a similar operation time without the assistance of the EP system.

### 3. The key point is the change in QRS morphology

When there is no EGM guidance, the operator mainly focuses on the QRS changes of captured LBB. During HBP, due to the long HV interval, obvious QRS changes can occur from the capture of the ventricular septum to the capture of the His bundle, which is easy to observe and judge. When PVC with RBBB morphology or paced RBBB pattern is noted during fixation, it can prompt the operator that the depth of screwing is close to LBB region. At this time, it is recommended to pause screwing in and perform a pacing test. During pacing in the LBB area, RBBB pacing pattern can appear in both LBBP and LVSP, it is necessary to observe the subtle changes between the two paced ECG morphologies to help differentiate them. These changes in QRS are reflected in the following points: (1) After LBB capture, there are two paced patterns: S-LBBP and NS-LBBP. The right bundle branch delay in S-LBBP is more pronounced due to the lack of peripheral myocardium capture, with a wider QRS complex and a deep S wave in lead V5. (2) When LBB is captured, the Stim-LVAT in lead V5 may be abruptly shortened by about 10ms. If the ECG monitoring cannot be measured, it can be repeatedly verified by changing the output voltage, and the shape and width of the QRS in the corresponding lead can be observed synchronously. The key point is to find differences in QRS changes for identification, and the S-LBBP pattern is the most sensitive and specific feature.

The techniques for repeated verification are recommended as follows: (1) Pacing with lower output (1V/0.5ms) first, and then pacing with higher output (10V/0.5ms), paying attention to changes in paced ECG morphology and Stim-LVAT. (2) Pacing test method: reduce the output in steps (can reduce 0.1V voltage or 0.1ms pulse width to adjust slightly output current) to distinguish the capture threshold of LV septal and LBB, or increase the pacing frequency to more than 130 bpm can also help differentiate due to the differences of refractory period of the myocardium and conduct bundle^12^.

### 4. Abrupt shortening of Stim-LVAT is important

In previous studies, a sudden shortening of Stim-LVAT during increasing output was taken as important evidence for the capture of LBB. In our study, abrupt shortening of Stim-LVAT is equally important, but since the four-channel ECG monitor does not have a measurement function, only obvious changes of Stim-LVAT can be observed with the naked eye.

Some studies proposed to use the V6-V1 interpeak interval to judge the capture of the LBB^18^, but in this study, since the V6-V1 interpeak interval could not be measured, this criterion was not applicable.

### 5. Patients in the CLBBB group were more likely to be identified and judged

LBBP in 100% of patients with CLBBB in this study cohort could be accurately judged intraoperatively. From the intrinsic LBBB morphology, to the narrow QRS of septal-derived PVC, and then to the typical RBBB morphology after LBB capture, sufficient and obvious mutations can be easily identified and judged. Activation from ventricle retrograde to atrium through the lesioned His bundle is rare in LBB block patients, so LBBP across the distal end of the block site is more likely to show S-LBBP patterns at low output, accounting for 94.7% (N=36/38) in this study.

### 6. The role of PSA

In addition to the routine pacing analysis during operation, PSA can also provide the recording function of EGM. It can help to record the conduction bundle potential with an amplitude greater than 0.1mv ahead of the V wave, and can record the potential COI in some cases (Figure 5). The value of the large potential with COI is to remind the operator that the lead tip is more accurately positioned in the conduction bundle. If the immediate capture threshold is too high, it is not necessary to screw deeply or change the position. It only takes several minutes to wait for the threshold to gradually decline after acute injury. Meanwhile, the observation of the amplitude change of V wave COI in PSA can help to determine whether the ventricular septum is perforated and avoid excessive screwing of lead^19^.

### 7. The value of LBB COI

Patients with LBB potential accompanied by potential COI showed a lower and stable capture threshold than patients without LBB potential COI. It was considered that patients with LBB potential COI had electrode tip closer to the left bundle branch, so a lower capture threshold could be obtained, and the threshold reduction was more obvious following the acute injury stage^20, 21^.

### 8. Study limitations

This study was completed by two experienced operators who have ample clinical experience in the judgment of LBB capture. Therefore, it is recommended that operators who are still in the learning curve first familiarize themselves with the electrophysiological characteristics and operating procedures of LBBP, and then transition to the simplified method.

This study is a single-center study with a sample size of 143 cases. Patients with acute anterior myocardial infarction, hypertrophic cardiomyopathy and nonspecific IVCD were not included in the group. Considering the difficulties in operation and judgment that may occur in patients with the above-mentioned basis, it is possible to expand the study to include the above population. This study did not involve other stylet-driven pacing leads, especially leads with non-conductive helical ends may have different implantation results.

## Conclusions

The simplified LBBP implantation method without an EP system and only relying on a simplified body surface ECG combined with PSA is clinically feasible and safe. It is recommended that the operator pay attention to the QRS mutation during LBBP procedure, including the S-LBBP/NS-LBBP pattern, and the QRS morphological changes accompanying with obvious changes of Stim-LVAT, which are key points that can be observed with the naked eye for judging the captured LBB. The new simplified method of judgement for LBB captured can also be extended during postoperative follow-up. At the present, there are many pacemaker implant centers do not have the sophisticated EP system, thus, a simplified method, especially an automated simplified method, should be pursued for promotion of LBBAP, a physiological pacing modality.

## Data Availability

The datasets used and analyzed during the current study available from the corresponding author on reasonable request.

## Nonstandard Abbreviations and Acronyms

LBBP: left bundle branch pacing
ECG: electrocardiogram
EP system: electrophysiology recording system
PSA: pacing system analyzer
CLBBB: complete left bundle branch block
CRBBB: complete right bundle branch block
NS-LBBP: nonselective left bundle branch pacing
S-LBBP: selective left bundle branch pacing
LVSP: left ventricular septal pacing
Stim-LVAT: time from stimulus to left ventricular activation
PVC: premature ventricular contraction
LBB: left bundle branch
COI: current of injury
LVEF: left ventricular ejection fraction.

## Acknowledgments

We thank Dr Xiaohong Zhou (Cardiac Rhythm Management, Medtronic Inc., Minneapolis, Minnesota, United States) for diligent review of this article.

## Sources of Funding

This work was supported by the Key Research and Development Program of Zhejiang (grant no. 2019C03012) and University-Industry Collaborative Education Program (grant no.22097154062650).

## Disclosures

None.

## What is known?

- LBBP has been shown to be effective and safe in patients with pacing indications.
- During LBBP, the right ventricle activation was relatively delayed when the LBB was captured, the QRS morphology would change correspondingly, and Stim-LVAT would be suddenly shortened. LBB potential and COI could be recorded in the LBB area, which could help confirm the LBB capture.

## What the study adds

- This study proposed a simplified LBBP implantation method without an EP system and only relying on a simplified body surface ECG combined with PSA.
- The novel LBBP implantation method does not increase implantation risk compared with the conventional implantation method, and is clinically feasible and safe.

## Notes

### Competing Interest Statement

The authors have declared no competing interest.

### Clinical Trial

ClinicalTrials.gov. NCT05553431

### Author Declarations

The approval of the Ethics Committee of the first affiliated hospital of Wenzhou Medical University

